# Feasibility, acceptability, and cognitive benefits of a theory-informed intervention to increase Mediterranean diet adherence and physical activity in older adults at risk of dementia: the MedEx-UK randomised controlled trial

**DOI:** 10.1101/2023.07.04.23292172

**Authors:** A Jennings, OM Shannon, R Gillings, V Lee, R Elsworthy, G Rao, S Hanson, W Hardeman, S-M Paddick, M Siervo, S Aldred, JC Mathers, M Hornberger, AM Minihane

## Abstract

**Background:** Despite an urgent need for multi-domain lifestyle interventions to reduce dementia risk there is a lack of interventions which are informed by theory- and evidence-based behaviour change strategies and no interventions in this domain have investigated the feasibility or effectiveness of behaviour change maintenance.

**Objectives:** We tested the feasibility, acceptability, and cognitive effects of MedEx-UK, a personalised theory-based 24-week intervention to improve Mediterranean-diet (MD) adherence alone, or in combination with physical activity (PA), in older-adults at risk of dementia, defined using a cardiovascular risk-score of >10%.

**Design:** 104 participants (74% female, 57–76 years) were randomised to three parallel intervention arms: 1) control, 2) MD, or 3) MD+PA for 24-weeks and invited to an additional 24-week follow-up period with no active intervention. Behaviour change was supported using personalised targets, a web-based intervention, group sessions and food provision.

**Results:** The intervention was feasible and acceptable with the intended number of ‘at risk’ participants completing the study. Participant engagement with group sessions and food provision components was high. There was improved MD adherence in the two MD groups compared with control at 24-weeks (3.7 points on a 14-point scale (95% CI 2.9, 4.5, p <0.01) and 48-weeks (2.7 points (95% CI 1.6, 3.7) p<0.01). Intervention did not change objectively measured PA. Improvements in general cognition (0.22 (95% CI 0.05, 0.35, p =0.01), memory (0.31 (95% CI 0.10, 0.51, p <0.01), and select cardiovascular outcomes captured as underpinning physiological mechanisms were observed in the MD groups at 24-weeks.

**Conclusion:** The intervention was successful in initiating and maintaining dietary behaviour change for up to 12-months, which was likely due to the intense early support provided which resulted in cognitive benefits. These results will inform larger-scale complex behaviour change interventions with health and well-being endpoints.

## Introduction

Dementia is a major public health concern with a substantial social and economic cost (1). Given the considerable and rising prevalence of this condition, the identification of feasible, acceptable and effective dementia prevention strategies is a major research priority (2). The modification of behavioural risk factors, including eating behaviours and physical activity, may play a key role in mitigating dementia risk (3).

Improving both dietary behaviours and physical activity (PA) levels could have additive and synergistic effects on brain health through overlapping physiological processes and activation of common intracellular pathways (4). Recent studies have reported beneficial effects on cognitive function in large-scale, multi-domain interventions which comprise dietary and PA changes (5) including those in specific ‘at risk’ sub-groups (6, 7).

Data from randomised controlled trials (RCTs) suggest that intervention to support adoption of a MD can improve cognitive function (8, 9). Our previous prospective cohort research indicated that a three-point increase in Mediterranean Diet Adherence Screener (MEDAS) score on a 14- or 15-point scale was associated with up to five less years of global cognitive ageing (10). Increased PA has also been associated with improved cognitive function and reduced dementia risk (11, 12). Several mechanisms could explain such effects, including improvement of cardiometabolic health (13), through physiological processes such as blood brain barrier function, cerebrovascular function, inflammation and β-amyloid clearance (14–16).

To our knowledge only one previous intervention has examined the impact of multidomain behaviours, including MD adoption, on neurocognitive function, with none conducted in the UK (17). This previous six-month MD plus exercise intervention improved spatial working memory performance in older adults in Australia (17). In addition, there are a lack of MD and PA interventions which are informed by theory- and evidence-based behaviour change strategies and no interventions in this domain have investigated the feasibility or effectiveness of behaviour change maintenance. Various barriers make adoption of a MD in a non-Mediterranean setting challenging, including cultural identity and acceptability of MD components, which necessitates careful intervention development (18). Development of theory- and evidence-based interventions and feasibility testing are critical phases in complex intervention development (19), hence the importance of establishing the feasibility of intervention with the MD in a UK setting prior to progression to a definitive RCT.

In the current manuscript, we report the primary and secondary outcomes of the MedEx-UK study, a 24-week multi-domain, theory-based intervention to improve MD adherence alone, or in combination with PA, in older adults at risk of dementia, defined using a cardiovascular risk-score (20). The intervention targets key influences on behaviour based on the Capability, Opportunity, Motivation and Behaviour Model (COM-B model) and includes evidence-based behaviour change techniques (BCTs) to encourage initial behaviour change and maintenance of any changes (21, 22). Following this 24-week intervention we invited all participants to a further 24-week follow-up period with no active intervention to investigate behaviour change maintenance. The primary outcomes were feasibility and acceptability of the intervention and the secondary outcomes included behaviour change (changes in MEDAS score and PA levels), cognitive function, cardiometabolic health (BMI and 24-hour ambulatory blood pressure) and process measures such as theory-based mediators of behaviour change. Data are presented for outcomes at both 24- and 48-weeks follow-up.

## Methods

### Study design

The study was pre-registered with ClinicalTrials.gov (NCT03673722) and the details of the protocol have been published (20). Briefly, participants from three UK centres (Norwich, Newcastle, and Birmingham) were randomised to a personalised, multi-domain intervention into one of three parallel intervention arms: 1) control, 2) MD, and 3) MD+PA. The main 24- week intervention took place between March 2019 and September 2020 and the 24 to 48- week trial add-on behaviour maintenance phase was completed by March 2021.

The sample size calculation for the study, based on dietary change of three-points on the MEDAS indicated 90 participants (30 participants in each arm) would be required to complete the study, which was increased to 108 participants to account for a 20% drop-out rate (20). With this sample size the smallest detectable change in MEDAS score with 90% power and 5% error was 1.23 points, within our minimum target for dietary change of 3 points, suggesting we would have a sufficiently precise estimate of whether a dietary change is achievable in this study.

### Participants

Individuals aged 55 to 74 years were recruited to take part in the intervention through primary care, in collaboration with the local Clinical Research Networks at each study site, and via direct-to-public advertisements. Full details of the study inclusion/exclusion criteria can be found elsewhere (20). Briefly, participants were required to: 1) have a QRISK2 score ≥10%, which indicates a ≥10% risk of having a cardiovascular event in the next 10 years; 2) have normal cognitive function as determined by a Montreal Cognitive Assessment (MoCA) score ≥ 23 (23); 3) be free of mild cognitive impairment, dementia or other severe neuropsychological complaints; 4) have a baseline MEDAS score <9 according to a modified version of MEDAS (24); and 5) undertake <90 minutes self-reported moderate-intensity PA each week. Eligibility to participate was determined through online, telephone and in-person screening sessions. Participants provided initial consent for the 24-week study intervention during the online screening, and written consent during the in-person screening. During the initial 24-week intervention, participants were invited to take part in the 24 to 48-week behaviour maintenance phase, and further written consent was obtained.

### Randomisation

Individuals who were deemed eligible to participate in MedEx-UK were allocated randomly to one of the three study intervention arms within each centre, with minimisation for MEDAS score (low = 0-4; high = 5-8) and sex, to ensure treatment arms were balanced for these parameters. Randomisation and allocation were completed by researchers who were not blinded to group assignment.

### Intervention phase

The first 24 weeks of the study comprised an intensive intervention period, during which participants in the MD and MD+PA arms were encouraged to change their behaviour via a combination of personalised targets based on participants’ self-assessment of their MD consumption and PA levels, a web-based intervention, group sessions with facilitators trained in behaviour change techniques, and supermarket vouchers or food delivery to support behaviour change and the potential extra costs of Mediterranean foods. Subsequently, participants were invited to take part in a behavioural maintenance phase (weeks 24-48), during which they had continued access to the web-based intervention only.

The intervention targets were to improve MEDAS scores by at least three points and increase levels of activity to 150 minutes of moderate, or 75 minutes of vigorous, activity per week. These targets were personalised: as part of the website intervention, participants were asked to self-assess their consumption of the Mediterranean diet and their PA levels. They then received personalised feedback and were encouraged to focus on specific behavioural changes from which they were expected to benefit. Participants were encouraged to select their own goals to meet these targets, which were introduced in a gradual process. Due to the negative associations between alcohol consumption and brain health, participants were not asked to increase their alcohol intake but if they consumed alcohol to switch the type of alcohol they consumed to wine, preferably red wine.

The web-based intervention was administered via an interactive, modular platform called LEAP^2^, as described elsewhere (20). LEAP^2^ included the ‘Eating Well’ module, designed to help participants increase their MEDAS score by providing real-time access to their score and details of the goals they were meeting, and facilitating participants to choose their own goals based on individual food preferences. Full details of the MEDAS targets are presented in **Supplemental Table 1.**

The ‘Moving More’ module (accessible only by participants in the MD+PA arm) was designed to help participants increase their PA. The module included a questionnaire to allow participants to determine their current PA levels and receive an award based on the level achieved (bronze (≥100 min of moderate or 50 min of vigorous-intensity PA per week), silver (≥120 min of moderate or 60 min of vigorous-intensity PA per week) or gold (≥150 min of moderate or 75 min of vigorous-intensity PA per week). Participants were encouraged to set a goal of moderate and/or vigorous activity in minutes per week and LEAP^2^ provided tailored PA suggestions based around participants preferences for cost, intensity, and type (group or individual) of exercise, and guided participants through overcoming key barriers associated with increasing PA levels.

In addition, LEAP^2^ included a diary feature to help participants plan meals and PA, and links to the study dietary assessment tool (Intake24) and the food provision element of the MedEx- UK study. Participants were encouraged to visit LEAP^2^ regularly throughout the 24-week intervention period.

Participants in the MD and MD+PA arms were invited to attend four group sessions (at weeks 0, 2, 4 and 12) that were designed to complement the web-based intervention. The group sessions were two hours for the MD group and 2.5 hours for the MD+PA group and comprised ∼6 participants and ∼6 supportive others (i.e., a friend or relative to provide social support). The group sessions were designed to target key influences on behaviour change based on the COM-B model (25) and incorporated evidence-based behaviour change techniques (BCTs) to encourage change and maintenance. Due to the COVID-19 pandemic, group sessions were conducted both in-person (prior to March 2020) and via videoconferencing software (after March 2020 during ‘lockdown’ periods). Participants were notified of their intervention group allocation at the start of their first group session.

Finally, participants in the MD and MD+PA groups were provided with £30 per week in vouchers for an online food retailer. Participants were encouraged to purchase foods that contributed to their MEDAS target score but this was not monitored. In cases where online food delivery was not possible (e.g., due to delivery restrictions to rural areas), participants were provided with equivalent vouchers for a supermarket of their choice.

Participants in the control group received dietary and PA advice in accordance with the UK National Institute for Health and Care Excellence (NICE) guidelines for individuals with a moderately elevated QRISK2 score (26). They also attended a one-hour group session at week 0, during which they were informed of their intervention group allocation and received a brief verbal presentation outlining the importance of a control group in research. Following completion of the 24-week intervention phase, the control group received £240 shopping vouchers (equivalent to £10 per week participation) as remuneration.

### Behavioural maintenance phase

Following the initial 24-week intervention period, consenting participants entered a behavioural maintenance phase during which they had continued access to the LEAP^2^ platform but no longer received group support sessions or food provision. The LEAP^2^ platform was modified to include content which aimed to support participants in maintaining healthy behaviour change achieved during the initial study intervention period, full details are provided in **Supplemental Methods 1**. This study maintenance phase was a trial add-on initiated after participants were recruited and consented to the main 24-week intervention.

### Outcomes

A baseline assessment was conducted in-person at a clinical testing facility prior to the study intervention, during which cognitive, vascular and biological outcomes were evaluated (1). The study protocol was adapted at the onset of the COVID-19 pandemic to minimise participant-researcher contact and to ensure safety and compliance with social distancing measures. Therefore, only a sub-set of secondary measurements, not including biological samples, or MRI were obtained at 24- and 48-weeks via remote (i.e., at-home) data collection. Specific adaptations have been highlighted for each measurement below, where relevant.

### Feasibility and acceptability

The feasibility of the intervention was assessed using recruitment and retention rates. Intervention fidelity and participant engagement were evaluated via group session attendance (intervention phase) and self-reported usage of LEAP^2^ ^(^intervention and behaviour maintenance phases) in the MD and MD+PA groups. Acceptability of the intervention was assessed at 24- and 48-weeks by a custom questionnaire using 5-point Likert-type scales, informed by the Theoretical Framework of Acceptability (27).

### Dietary assessment

Dietary intake, to determine level of adherence to the MD, was evaluated via two different approaches. Firstly, participants completed an online version of the 14-point MEDAS questionnaire (24), which was the primary dietary outcome measure in this study. Secondly, participants completed a series of 24-hour recalls (on five non-consecutive days at baseline and at 24- and 48-weeks) via Intake24, a validated online dietary assessment tool (28). These data were also used to calculate adherence to the 14-point MEDAS scale as detailed in **Supplemental Table 1**.

### Physical activity

PA levels were recorded for all participants throughout the entire intervention and behaviour maintenance periods via wrist worn activity monitors (Vivosmart 3, Garmin). The activity monitors were set to show the time and date only, to prevent participants receiving any activity-based feedback. Age, height, and weight were entered when setting up the devices to improve accuracy. The devices recorded total step count, heart rate and PA energy expenditure. In addition, total activity levels in minutes of moderate intensity PA per week were calculated as: moderate minutes (defined as 40–59% heart rate reserve) + (vigorous minutes (≥ 60% heart rate reserve)*2) (29).

### Cognitive function

Cognitive function was determined using an extended version of the neuropsychological test battery (NTB)(20) measured at baseline, 24- and 48-weeks (**Supplemental Methods 2**). Additionally, we included assessments of spatial navigation via the virtual reality Supermarket Trolley Task(30) and the Sea Hero Quest Test(31). The duration of each cognitive assessment was approximately 90 minutes. Baseline assessments were conducted in-person at a clinical testing facility and follow-up assessments at 24- and 48-weeks were conducted remotely via video conferencing software, to reduce in-person contact while COVID-19 social distancing measures were in effect. A researcher was present virtually during the testing and paper-based cognitive tests were posted to participants before the session. It was not possible to collect data on the spatial navigation tasks during these remote sessions.

Scores from each test were converted to Z scores standardised on baseline grand mean and standard deviation with response time variables reversed by [Z * -1], so higher scores always suggested better outcomes. Individual Z scores test scores were mean aggregated into summary scores for: Processing speed [Digit symbol substitution (total correct); Trail Making Test (A, seconds)]; Executive Function [Controlled Oral Word Association Test (total); Categorical verbal fluency test (total); Trail Making Test (B-A, seconds); Wechsler Memory Digit Span (backwards, total)] and Memory [Visual paired (immediate and delayed totals); Verbal paired (immediate and delayed totals); Rey Auditory Verbal Learning Test (immediate and recall)]. A general cognition score was calculated using all Processing Speed, Executive Function and Memory tests.

### BMI

At baseline, height and weight were measured after an overnight fast by a member of the research team using standard laboratory techniques and used to calculate body mass index (BMI). At 24- and 48-weeks, due to social distancing measures during the COVID-19 pandemic, participants were asked to measure their body weight at home using either their own electronic scales or those provided by the research team.

### 24-hour ambulatory BP

24-hour ambulatory BP (AMBP) was measured at baseline and week-24 using portable devices (Mobil-O-Graph, Stolberg, Germany and Spacelab Healthcare, Washington, United States) which consisted of an inflatable cuff attached to a small monitoring system. The cuff was secured around the upper arm and readings were taken every 20 minutes during daytime (06:00 to 22:00) and every hour overnight (22:00 to 06:00) for an entire 24-hour period.

### Process evaluation

The process evaluation was informed by UK Medical Research Council guidance for process evaluation (32). Here we present the quantitative measures related to mechanism of impact. The findings from interviews with group session facilitators and focus groups with participants, which focus additionally on contextual factors and implementation (e.g., fidelity) will be reported separately. Hypothesised mediators of behaviour change (intention, perceived control and self-reported use of behaviour change techniques) were assessed in all groups at baseline (intention and perceived control only), 24- and 48-weeks using five-point Likert-type scales.

### Statistical analyses

Between group differences in group-session attendance and use of the online-platform were examined using a 2-sample t-test or χ² test for categorical data. The effect of the intervention on eating behaviour and other outcomes at 24- and 48-weeks were assessed using ANCOVA, with the 24-week value as the dependent variable, group as the independent variable. We compared the difference in the mean of the control group with the mean of the two MD intervention groups (contrast 1), and the mean of the MD and MD+PA groups (contrast 2). Covariates included baseline value, study site, and baseline BMI. The cognitive outcomes were additionally adjusted for age and years of education. We checked for effect modification by sex by including an interaction term for group*sex in the models. Data are presented as the difference in mean values at 24- or 48-weeks for the two intervention groups (mean MD+PA and MD) minus the control group.

For eating behaviour change, we also examined if change over 24-weeks was associated with maintenance or further change at 48-weeks by including an interaction term for group*continuing to 48-weeks (y/n) in the model. We also calculated the percentage of participants who changed their diets sufficiently to meet the criteria for individual MEDAS components at 24-weeks. Finally, participants across all three groups (control, MD+PA, MD) were assigned to tertiles of 24-week change in MEDAS score and minutes of moderate activity and associations with cognitive and cardiometabolic outcomes at 24-weeks were examined.

All data are presented as unadjusted mean (SD) at individual timepoints, change (95% CI) or percentages where indicated. All analyses were performed using STATA (version 16; StataCorp).

## Results

### Feasibility and acceptability

Of the n=2776 participants who completed online screening, n=239 met the criteria and attended in-person screening, n=104 (74% female, 57–76 years (mean 67.4 years (SD 4.6)) were recruited to the MedEx-UK study between 15^th^ April 2019 and 10^th^ January 2020. The main 24-week intervention was completed by n=99 (5% drop out rate) of whom n=76 (77 %) consented and n=69 (9% drop out rate) completed to the 24 to 48-week trial add-on behaviour maintenance phase (**Supplementary Figure 1 and Table 1**). Complete data for the change in dietary and physical activity behaviour, were available for n=87 completers (88%) at 24-weeks and n=52 completers (75%) at 48-weeks.

**Table 1:** Baseline characteristics of the MedEx-UK study participants, according to intervention group

| **Characteristic** | **MD+PA (n=35)** | **MD (n=35)** | **Control (n=34)** |
| --- | --- | --- | --- |
| Sex, female | 25 (71.4%) | 27 (77.1%) | 25 (73.5%) |
| Age, years | 68.1 (5.1) | 67.3 (4.3) | 67.1 (4.4) |
| Ethnicity, white | 34 (100%) | 35 (100%) | 33 (97%) |
| IMD, decile | 5.9 (3.1) | 5.2 (2.6) | 6.3 (2.3) |
| BMI, kg/m<sup>2</sup> | 27.5 (4.2) | 30.1 (5.0) | 28.4 (3.5) |
| MEDAS, score | 6.8 (2.2) | 5.9 (2.0) | 6.8 (2.1) |
| Moderate activity, min/d | 181 (154) | 230 (205) | 261 (308) |
| QRISK, score | 17.2 (5.8) | 16.7 (4.9) | 15.9 (4.7) |
| General cognition score, z-score | -0.07 (0.5) | 0.12 (0.6) | -0.01 (0.6) |
| Processing speed score, z-score | -0.14 (0.8) | 0.09 (0.9) | 0.05 (0.9) |
| Executive function score, z-score | -0.12 (0.6) | 0.13 (0.8) | -0.01 (0.6) |
| Memory score, z-score | 0.05 (0.7) | 0.05 (0.6) | -0.08 (0.7) |
| 24hr mean systolic BP, mm Hg | 128 (12.5) | 127 (14.3) | 128 (11.1) |
| 24hr mean diastolic BP, mm Hg | 78.3 (10.3) | 77.0 (9.6) | 75.2 (9.8) |
| 24hr mean pulse pressure, mm Hg | 51.2 (9.3) | 52.2 (10.9) | 53.0 (7.5) |
| 24h systolic BP variability, mm Hg | 10.8 (2.9) | 10.6 (2.5) | 10.1 (2.9) |
| 24h diastolic BP variability, mm Hg | 11.0 (3.7) | 11.0 (4.3) | 10.8 (3.5) |
| 24h pulse pressure variability, mm Hg | 16.3 (11.9) | 13.9 (8.9) | 14.1 (8.0) |
| Ambulatory Arterial Stiffness Index | 0.61 (0.18) | 0.58 (0.19) | 0.51 (0.29) |

Values are mean (SD) or n= (%). Data was missing for BMI (n=1 MD+PA), IMD (n=1 MD+PA; n= 1 MD), Moderate activity (n=1 MD+PA; n=2 Control) and QRISK (n=4 MD+PA; n= 3 MD; n=5 Control). IMD = Index of Multiple Deprivation, MD = Mediterranean diet, MEDAS = Mediterranean Diet Adherence Screener, PA = Physical activity, QRISK = cardiovascular risk score.

Engagement with the group sessions was high, with participants attending 3.6 (SD 0.7) and 3.5 (SD 0.9) of the four group sessions in the MD+PA and PA groups, respectively (**Supplemental Table 2**). Most participants (84%) reported accessing the online platform once per month or less, with the average session length 15 to 30 minutes. Uptake of food delivery or supermarket vouchers was 100% each month. Ninety-five percent of participants reported the intervention to be acceptable, with no significant difference between the MD and MD+PA intervention groups (**Supplementary Figure 2**). Overall, participants rated the acceptability of the online platform lower than other intervention components (**Supplemental Table 3**). Participants rated their understanding of how the intervention aimed to facilitate behaviour change highly and reported a good fit with their beliefs about behaviour change (both average scores 4.3 out of possible five, **Supplemental Table 3**).

### Eating behaviour

After the 24-week intervention, there was improved MD adherence in the two MD groups compared with control when assessed using the MEDAS questionnaire (3.7 points (95% CI 2.9, 4.5, p <0.01) **Figure 1A and Supplemental Table 4**) and using 24-hour recall (3.4 points (95% CI 2.4, 4.4, p <0.01), (**Figure 1B and Supplemental Table 5**). There was no evidence of a group by sex interaction (data not shown). Likewise, at 48-weeks there was improved adherence in the two MD groups compared with control when assessed using the MEDAS questionnaire (2.7 points (95% CI 1.6, 3.7) p <0.01) and using 24-hour recall (2.6 points (95% CI 1.5, 3.8) p <0.01) data (**Figures 2A and 2B**). Participants in the MD group who participated in the maintenance phase had significantly higher MEDAS scores at 24- weeks compared to those who did not continue (between group difference of 1.5 points (95% CI 0.4, 2.8) p=0.01), with no significant difference in the MD+PA group (between group difference of 0.9 points (95% CI -0.4, 2.3) p=0.17).

**Figure 1:**
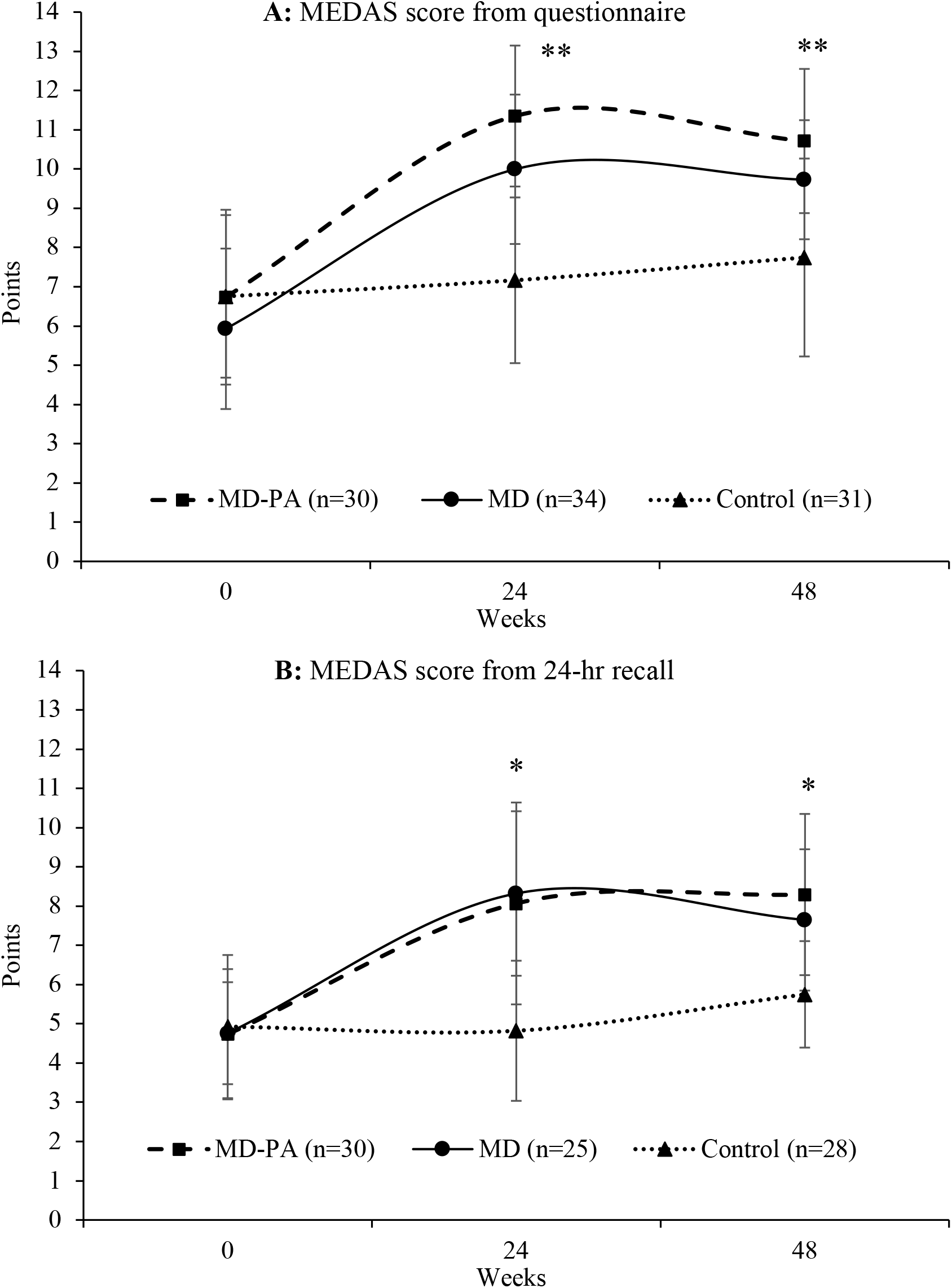
Mediterranean Diet Adherence Screener (MEDAS) score by intervention group at baseline, 24- and 48- weeks calculated by questionnaire and 24-hour recall. Values represent unadjusted means (SD) from MEDAS questionnaire (panel A) and 24-hour recall (panel B). P-value for group and contrast 1 (Control v. (MD + MD+PA)) *< 0.01 or ** <0.05 at relevant time point compared to baseline, calculated using ANCOVA (adjusted for baseline value, study site and baseline BMI). P-values for contrast 2 (MD v. MD+PA) were non-significant at all timepoints compared to baseline as were all contrasts comparing values at 48- to 24-weeks. Participant numbers at 48-weeks were n=20 MD+PA, n=22 MD, n=21 Control for questionnaire data and n=17 MD+PA, n=17 MD, n=12 Control for 24=hr recall data. MD = Mediterranean diet, MEDAS =Mediterranean Diet Adherence Screener, PA = Physical activity.

**Figure 2:**
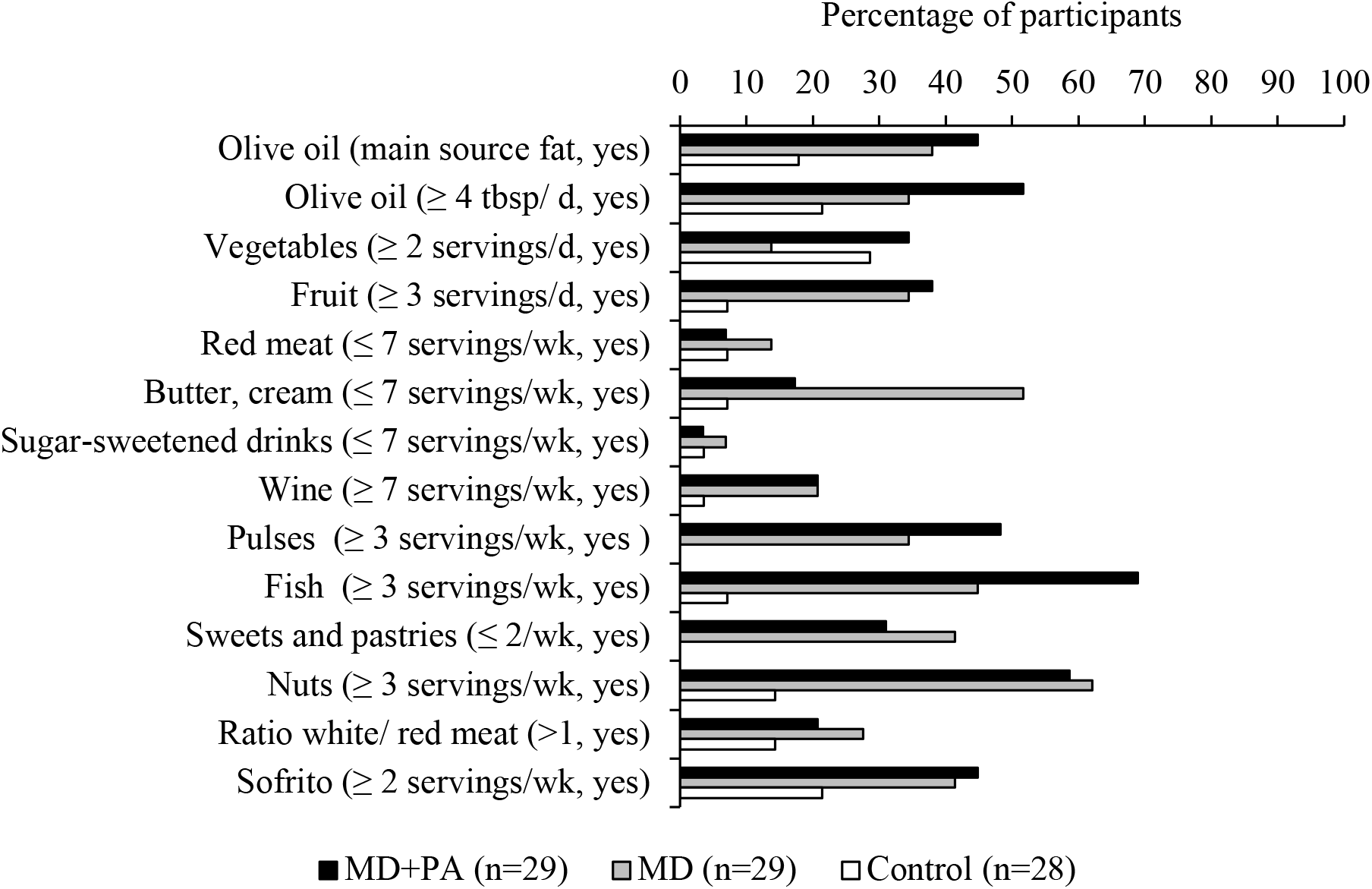
Proportion of participants adapting to meet the criteria for individual Mediterranean Diet Adherence Screener components at 24-weeks by intervention group in 86 MedEx-UK participants. Bars represent the percentage of participants who met the criteria at 24-weeks but not at baseline according to the questionnaire data. Only participants with complete data for all components were included (n=86).

Consumption of all components of the MEDAS score improved in the MD diet groups compared with control over the 24-week intervention except for vegetables and sugar-sweetened drinks (when assessed by questionnaire), and sugar-sweetened drinks and butter and cream (when assessed by 24-hour recall) (**Supplemental Tables 4-5**). According to the MEDAS questionnaire data, red meat and sugar-sweetened beverages were the recommendations met by the highest proportion of participants at baseline (**Supplementary Figure 3)** and the nut and fish components were most likely to be changed by participants in the MD groups, with 60 % and 57 % of participants, respectively, adapting their diet sufficiently to meet the recommendations over the 24-week intervention (**Figure 2**). Using 24-hour recall data, the ratio of white to red meat and sofrito were the components that were most likely to be adapted (**Supplementary Figure 4**).

### Physical activity

Total number of steps, energy expenditure and minutes of moderate activity increased in the MD+PA group and decreased in the MD and control groups after the 24-week intervention, but no significant between-group differences were observed (**Table 2**). Likewise, at 48-weeks no significant between-group differences were observed in PA (**Table 2**). There was no evidence of a group by sex interaction for minutes of moderate activity at 24-weeks and there was no difference in minutes of moderate activity at 24-weeks between the participants in the MD+PA group who did, or did not, continue to 48-weeks (data not shown).

**Table 2:** Physical activity and cardiometabolic outcomes at baseline, 24- and 48-weeks by intervention group in 99 MedEx-UK participants.

|  | MD+PA |  |  | MD |  |  | Control |  |  | P1 | P2 | P3 |
| --- | --- | --- | --- | --- | --- | --- | --- | --- | --- | --- | --- | --- |
|  | n= | Baseline | 24-weeks | n= | Baseline | 24-weeks | n= | Baseline | 24-weeks |  |  |  |
| Total Steps, d | 28 | 5743(2100) | 6348(3343) | 31 | 6137 (2364) | 5760 (2501) | 31 | 6282(2562) | 5913(2680) | 0.13 | 0.23 | 0.31 |
| Energy expenditure, kcal/week | 28 | 281 (171) | 327 (304) | 31 | 322 (189) | 277 (154) | 30 | 303 (189) | 239 (131) | 0.06 | 0.12 | 0.30 |
| Moderate activity, min/week | 28 | 191 (163) | 262 (183) | 31 | 233 (214) | 228 (195) | 31 | 256 (311) | 213 (210) | 0.10 | 0.14 | 0.49 |
| BMI, kg/m<sup>2</sup> | 33 | 27.6 (4.2) | 27.1 (4.4) | 34 | 30.3 (5.0) | 30.3 (5.2) | 32 | 28.4 (3.6) | 28.3 (3.4) | 0.20 | 0.67 | 0.06 |
| 24hr mean SBP, mm Hg | 13 | 128 (14.0) | 125 (14.4) | 13 | 128 (14.2) | 130 (15.5) | 11 | 131 (9.8) | 128 (10.6) | 0.57 | 0.58 | 0.04 |
| 24hr mean DBP, mm Hg | 13 | 75.7 (10.0) | 75.4 (10.1) | 13 | 80.2 (10.7) | 75.8 (6.0) | 11 | 76.4 (7.5) | 75.1 (4.8) | 0.89 | 0.67 | 0.66 |
| 24hr mean PP, mm Hg | 13 | 52.4 (7.3) | 51.4 (7.2) | 13 | 51.4 (11.0) | 53.7 (12.1) | 11 | 54.7 (7.8) | 53.1 (8.2) | 0.87 | 0.37 | 0.08 |
| 24h SBP CV, mm Hg | 13 | 10.5 (2.7) | 9.5 (1.8) | 13 | 9.6 (1.5) | 9.6 (1.8) | 11 | 9.4 (1.9) | 9.6 (1.8) | 0.32 | 0.49 | 0.41 |
| 24h DBP CV, mm Hg | 13 | 10.2 (3.2) | 9.8 (3.6) | 13 | 9.5 (2.6) | 10.4 (2.7) | 11 | 10.6 (2.9) | 10.6 (2.6) | 0.32 | 0.85 | 0.10 |
| 24h PP CV, mm Hg | 13 | 14.0 (9.9) | 11.4 (6.9) | 13 | 15.6 (8.3) | 15.7 (9.1) | 11 | 14.1 (7.4) | 15.9 (8.4) | <0.01 | 0.02 | 0.02 |
| AASI | 13 | 0.58 (0.19) | 0.50 (0.10) | 13 | 0.62 (0.17) | 0.64 (0.16) | 11 | 0.60 (0.20) | 0.64 (0.14) | 0.01 | 0.13 | <0.01 |
|  |  |  | **48-weeks** |  |  | **48-weeks** |  |  | **48-weeks** |  |  |  |
| Total Steps, d | 17 | 5497(2187) | 5470(2647) | 21 | 6264 (2055) | 5683 (2499) | 18 | 5895(2284) | 5586(2822) | 0.85 | 0.81 | 0.96 |
| Energy expenditure, kcal/week | 17 | 228 (97) | 242 (134) | 21 | 310 (165) | 289 (201) | 16 | 301 (166) | 248 (195) | 0.56 | 0.66 | 0.67 |
| Moderate activity, min/week | 17 | 204 (167) | 246 (247) | 21 | 216 (209) | 314 (228) | 18 | 273 (316) | 288 (374) | 0.59 | 0.37 | 0.61 |
| BMI, kg/m<sup>2</sup> | 24 | 27.0 (3.2) | 26.3 (3.2) | 23 | 30.3 (4.7) | 29.9 (4.9) | 21 | 27.8 (3.2) | 28.0 (3.2) | <0.01 | 0.02 | 0.31 |

Values are unadjusted means (SD). P1 = p-value for group using ANCOVA (adjusted for baseline value, study site and baseline BMI), P2 = p-value for contrast 1: Control v. (MD + MDPA), P3 = p-value for contrast 2: MD v. MDPA. Moderate activity minutes have been derived from light, moderate and vigorous intensity minutes normalised to moderate intensity. AASI = Ambulatory stiffness index, CV = variability, DBP = diastolic blood pressure, MD = Mediterranean diet, PA = physical activity, PP = pulse pressure, SBP = systolic blood pressure.

### Cognitive function

After the 24-week intervention, there were improvements in test scores for general cognition (0.22 (95% CI 0.05, 0.35, p =0.01) and memory (0.31 (95% CI 0.10, 0.51, p <0.01) domains in the two MD groups compared with control **(Figure 3 and Supplemental Table 6).** These changes were determined by improvements observed in the Verbal Paired Associates task, a measure of verbal memory (4.2 (95% CI 0.06, 0.77, p <0.01, **Supplemental Table 7**). There were no significant differences in the test score for the processing speed domain, and the z-score for Executive Function was of borderline significance (p=0.05). Differences between the MD+PA and MD groups were not observed for any of the domains. At 48-weeks no between-group differences were observed in test scores in any domain.

**Figure 3:**
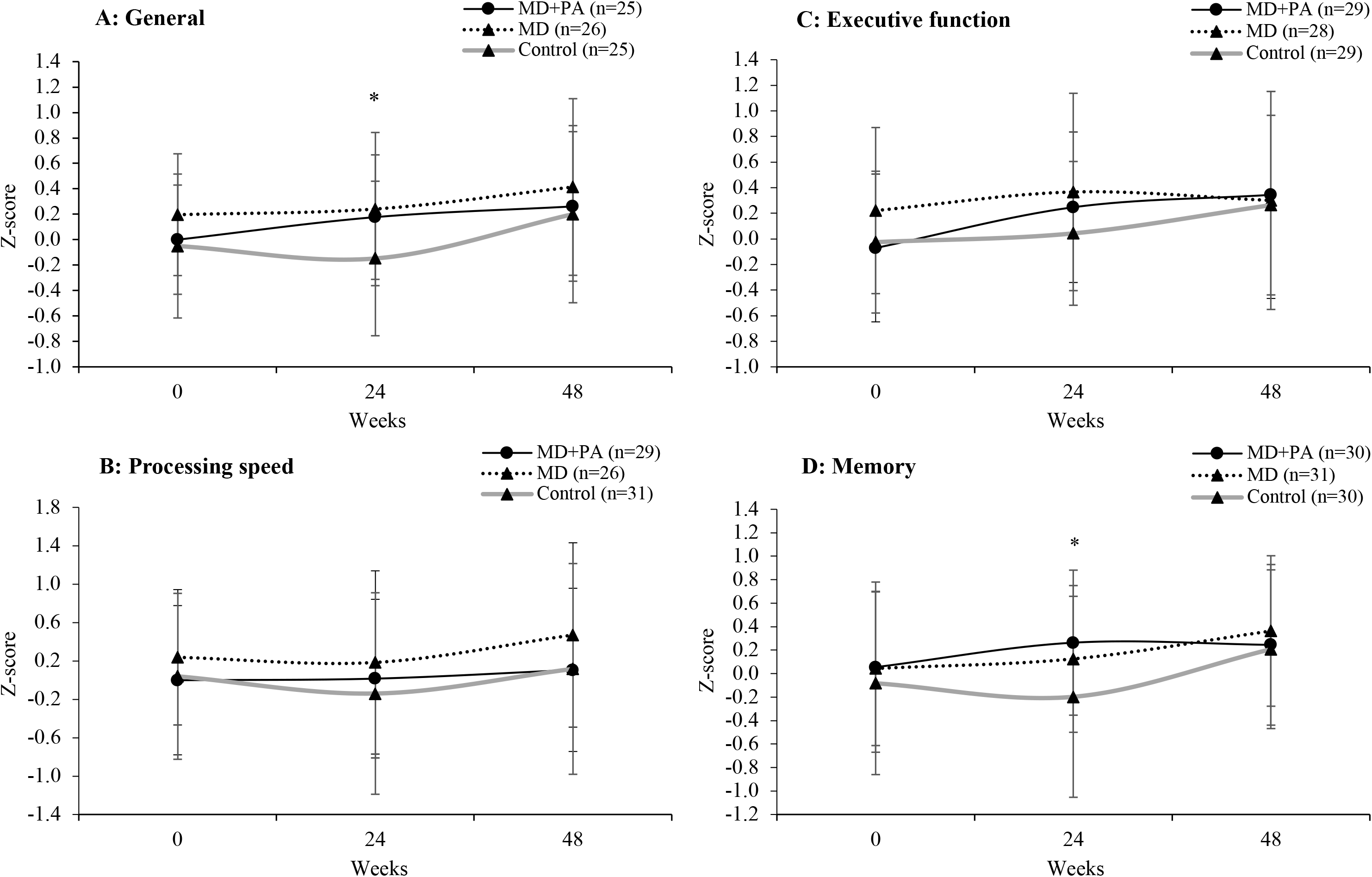
Cognitive summary scores by intervention group at baseline, 24- and 48- weeks. Values represent unadjusted means (SD) for General cognition (panel A), processing speed (panel B), executive function (panel C) and executive function (panel D). * P-value < 0.01 for group and contrast 1 (Control v. (MD & MD+PA)) at relevant time point compared to baseline, calculated using ANCOVA (adjusted for baseline value, study site baseline age, and years of education). P-values for contrast 2 (MD v. MD+PA) were non-significant at all timepoints compared to baseline as were all contrasts comparing values at 48- to 24-weeks. Missing data at 48-weeks were General cognition (MD+PA n=7, MD n=3, Control n=8), processing (MD+PA n=5, MD n=5, Control n=9), executive function (MD+PA n=6, MD n=10, Control n=11) and executive function (MD+PA n=11, MD n=9, Control n=8). Individual test scores were converted to Z scores standardised on Baseline grand mean and standard deviation with response time variables reversed by [Z * -1], so a higher time indicates a better outcome. Individual Z scores test scores were mean aggregated into Summary Scores for: Processing speed [Digit symbol substitution (total correct); Trail Making Test (A, seconds)]; Executive Function [Controlled Oral Word Association Test (total); Categorical verbal fluency test (total); Trail Making Test (B-A, seconds); Wechsler Memory Digit Span (backwards, total)] and Memory [Verbal paired immediate (total); Visual paired immediate (total); Verbal paired delayed (total); Visual paired delayed (total); Rey Auditory Verbal Learning Test (immediate); Rey Auditory Verbal Learning Test (recall)]. A general cognition score was calculated using all Processing speed, Executive function and Memory tests. MD = Mediterranean diet, MEDAS =Mediterranean Diet Adherence Screener, PA = Physical activity. Full data is presented in Supplemental Table 6.

In the Hayling test of Executive Function (another measure of executive function, not in the NTB test battery), response times reduced for section A (-5.0 seconds (95% CI -7.5, -2.5), p<0.01) and section B (-13.5 seconds (95% CI -21.8, -5.3), p<0.01) over the 24-week intervention in the two MD groups compared with the control group, with no differences observed in the number of errors made (raw or scaled) **(Supplemental Table 8)**. There were no significant differences in response times or number of errors made between-groups at 48- weeks **(Supplemental Table 8).** The Hayling overall scaled score improved in the two intervention groups relative to control at 24-weeks (0.7 (95% CI 0.4, 1.0), p<0.01) but not 48-weeks **(Supplemental Table 8).** Differences between the MD+PA and MD groups were not observed for any of the Hayling outcome measures.

Improvements in test scores in the general cognition (T3-T1 0.29 (95% CI 0.08, 0.50, p<0.01) and memory domains (T3-T1 0.35 (95% CI 0.10, 0.60, p<0.01) over 24-weeks were greater in the participants with the highest change in MEDAS score over the same time **(Supplemental Table 9).** Likewise, there were also improvements in the Hayling test scores with overall scaled scores improved in the participants with the highest change in MEDAS score (T3-T1 1.1 (95% CI -0.3, 1.9), p<0.01). For physical activity, fewer section B errors were observed in the participants with the greatest increases in moderate activity (T3-T1 1.8 (95% CI -3.3, -0.2), p=0.02, **Supplemental Table 10**).

### Cardiometabolic outcomes

At 24-weeks there was no significant intervention effect on BMI (**Table 2**) but at 48-weeks BMI reduced (-0.71 kg/m^2^ (95% CI -1.30, -0.13), p=0.02) in the MD+PA and MD groups compared with control. There were no intervention effects on 24-hour mean systolic, diastolic or pulse pressure at 24-weeks but a reduction in pulse pressure variability (-2.9 mm Hg (95% CI -5.3, -0.5), p<0.01) and Ambulatory Arterial Stiffness Index (-0.07 (95% CI – 0.2, 0.02), p=0.01) was observed in the MD+PA group. AMBP data were not collected at 48- weeks.

Improvements in pulse pressure variability (T3-T1 -3.4 (95% CI -6.1, -0.7), p=0.01, **Supplemental Table 9)** and Ambulatory Arterial Stiffness Index (T3-T1 -0.2 (95% CI -0.3, - 0.1), p<0.01) over 24-weeks were greater in those participants with the highest change in MEDAS score. No associations were observed between change in physical activity and cardiometabolic outcomes **(Supplemental Table 10**).

### Mechanism of impact measures

At baseline, participants were confident (perceived control) and motivated to change their diet and increase physical activity. Confidence and motivation reduced over the 24-week intervention period in all groups, with no significant between-group differences observed (**Supplemental Table 11**). Self-reported use of behaviour change techniques taught in the intervention was higher among intervention participants than control participants. Goal setting and incorporating dietary and physical activity change into daily routines were the most frequently utilised behaviour change techniques by intervention participants (with slightly lower levels for PA compared with diet). Conversely, social support and self-rewards were used least often (**Supplemental Table 12**).

## Discussion

This 24-week multi-domain, theory-informed intervention in older, ‘at risk’ adults living in the UK proved to be feasible and acceptable as judged by our ability to recruit the intended number of completers (with pre-specified characteristics), the high levels of retention at follow up and our ability to deliver the intervention as intended despite COVID-19 and social distancing restrictions. In addition, participant engagement with two of the three intervention components designed to support behaviour change was high, specifically the group sessions and uptake of food provision. Conversely, whilst the website was accessible to all participants, use was low and rated poorly. Focus groups highlighted this was mainly due to the poor functionality, which will need to be optimised prior to large-scale evaluation. Adherence to intervention was associated with cognitive (global cognition and memory) and cardiovascular (pulse pressure variability and arterial stiffness) benefits.

A previous study in the UK evaluating the feasibility of a peer support intervention to encourage adoption of a MD reported challenges with recruitment and retention of participants (33, 34). The successful recruitment and retention in the current study may be due to the use of individual-level recruitment, rather than the group-based approaches employed in the previous study, which may have ensured the inclusion of more engaged participants. The addition of a food provision component to remove barriers associated with the perceived higher price and inconvenience of healthy foods is also likely to have improved retention. Previous studies have shown that financial support improves adherence to a MD when accompanied by an educational intervention (35). We provided participants with options to choose MD components that met their personal food preferences, rather than being prespecified by study design, as personalisation has shown to lead to sustained changes in dietary behaviour (36).

The MedEx-UK intervention was also successful in improving eating behaviour with these changes maintained during the six months follow-up. A-priori we specified successful behaviour change as a three-point increase on the MEDAS. Participants in the intervention groups achieved a 3.7-point increase in MEDAS at 24-weeks, with a 2.7-point increase maintained at 48-weeks follow-up. Change in MEDAS scores of this magnitude are likely to be biologically and clinically important with previous studies reporting an approximate 30 % reduced risk of major cardiovascular events (16), a 12.6 to 20.7% reduced risk of dementia (37) and up to five years of reduced global cognitive ageing (10) with a change in MEDAS score of 2 -3 points.

Participants in the current study reported improved adherence to all dietary components (in particular, fish, nuts and olive oil), except for sugar-sweetened beverages which were habitual relatively low at baseline. Conversely, in the PREDIMED study, conducted in a Mediterranean-region, dietary changes were only apparent for foods attributable to the free products provided (olive oil and nuts), legumes and fish (16). This suggests that more food changes were required by UK participants to align with the MD, but these changes were achievable and maintained for one year.

Significant between group differences in PA were not observed and increases in the MD+PA group were modest at 24-weeks. These modest changes in PA of approximately 70 minutes per week were not entirely unexpected given that the study took place during COVID-19 lockdowns where activity opportunities were restricted. It was of interest that activity levels only increased in the MD+PA group and decreased in the other groups which may suggest that the PA component was effective at maintaining activity levels during COVID-19 lockdowns. A recent large US cohort study reported that increasing activity by 10 minutes per day could reduce preventable deaths by seven percent per year (38) and 10 minute activity bouts have been linked to improved cognition (39) suggesting that even small changes in activity are important from a public health perspective. Of note, at screening, all participants self-reported <90 minutes moderate-intensity PA each week, although at baseline, using directly measured activity, mean moderate activity was 191 minutes per week in the MD+PA group, with only 34 % of the group below the 90 minutes threshold. This highlights the weaknesses of subjective versus objective PA assessment and suggests refinement of PA methodology at screening is required in future studies to ensure recruitment of intended participants. This may be another factor to explain the moderate changes in PA we observed in the intervention.

Whilst the current study did not observe significant changes in PA, it is notable that the additional behaviour targets in the MD+PA group was not a deterrent to improving eating behaviour and was associated with improvements in cognition and cardiometabolic health. Pulse pressure and ambulatory stiffness index, measured using 24-hour ambulatory blood pressure, were improved in the MD+PA group alone at 24-weeks. We also observed a dose effect with greater improvements in cognition and cardiovascular health in participants with the highest levels of behaviour change. Research suggests there are synergistic associations between an individual’s lifestyle risk behaviours and health outcomes which highlights the importance of developing interventions that tackle multiple behavioural risk factors (40).

The intervention was effective at improving general cognition and memory (including verbal memory) over 24-weeks. It was unexpected that these improvements were not maintained at 48-weeks and may indicate that a longer duration of intervention is required for sustained cognitive benefits associated with modest maintained behaviour change. Our findings of improvements in general cognition and memory support those of the FINGER trial with the trend for the same beneficial effects on executive function (with exception of significant effect in the Hayling test), which again may reflect the shorter duration of our intervention (5). The Hayling processing speed component appeared to be the most sensitive cognitive measure and may be important to explore in future studies.

Process evaluation was an essential part of this feasibility study and provides us with important insights to inform the development of a larger-scale trial. The intervention recruited a highly motivated sample; participants reported high confidence and motivation to change their diet and increase physical activity. However, confidence and motivation reduced over the 24-week period. This may be due to unrealistic optimism at baseline, with participants becoming more realistic over time due to experiences with behaviour change (41). In addition, the COVID-19 lockdown and other restrictions may have made behaviour change especially challenging. The behaviour change observed after the intervention was not due to increased motivation and confidence and is more likely due to increasing participants’ use of behaviour change techniques, promoted by the intervention, in their daily lives. The most frequently used techniques (goal setting, building routines) facilitate behaviour maintenance. Participants in the MD+PA reported using these techniques slightly less frequently for PA, than for dietary change, which may have contributed to the differences between observed change in MD and in PA at 24 weeks. In contrast, social support was used least often. Although it was included in the group sessions, the restrictions resulting from the pandemic limited opportunities for social support, especially face-to-face.

Strengths of the current study include: i) the development of intervention components which targeted key influences on behaviour based on the COM-B model and included evidence-based behaviour change techniques, ii) the robust measurements of feasibility and acceptability, which were informed by Medical Research Council guidance for process evaluation (19), iii) the use of validated measurement tools for assessing the primary (diet and PA) and secondary (cognition and cardiometabolic) outcomes and iv) the inclusion of both behavioural and clinical data. The COVID-19 pandemic created unique challenges for the intervention study and restricted our ability to collect the data for our secondary cardiometabolic outcomes. However, our successful experiences around remote delivery of this complex intervention will be invaluable for the design and delivery of future interventions. We defined participants ‘at risk’ of dementia using a scale to monitor risk of cardiovascular disease which does not include assessment of other important risk factors for dementia, including cognitive function and family history. We were not successful in recruiting socio-economically disadvantaged and racially and ethnically diverse participants and the recruitment protocol and study design for a future study will need to be modified to ensure such inclusion. Further learnings for the development of a larger scale trial include the need to improve the poor conversion rate from screening to recruitment and to further develop and test intervention tools that are acceptable, feasible and inclusive to our target population, in particular, the online platform.

In conclusion, this feasibility study to increase Mediterranean diet adherence and physical activity in older adults at risk of dementia was acceptable and feasible. In addition, it was effective at improving eating behaviour, alone and when increased PA was an additional behavioural target. The intervention was also successful in behaviour change maintenance for up to 12 months, which was likely due to intense early support and investment to achieving long-term behaviour change. The behaviour change was associated with cognitive and cardiovascular benefits especially in the combined Mediterranean-style diet and physical activity intervention group. This feasibility testing will be essential in developing a larger scale intervention based on MedEx-UK and to other researchers in planning complex behaviour change interventions.

## Data Availability

The data that support the findings of this study are available from the corresponding author, upon reasonable request.

## Author contributions

SH, WH, MH, SMP, MS, SA, JCM and AMM were responsible for the conception and design of the work and funding acquisition. AJ, OS, RG, VL, RE and GR conducted the investigation. AJ accessed and verified the data and completed the statistical analyses. All authors had access to the study data and contributed to the data interpretation. AJ and OS wrote the original draft. All authors provided critical revision of the manuscript and approved it prior to publication.

## Acknowledgements

The authors would like to thank the participants of the MedEx-UK study.

